# Translational Study of using FOCM/TS Metabolites for Supporting Autism Spectrum Disorder Diagnosis

**DOI:** 10.64898/2026.01.06.26343525

**Authors:** Halil Arici, Marie Causey, Soma Patra, Uwe Kruger, Cristopher Antonio Villegas Uribe, Raun Melmed, Craig Ciuk, Sophia Crisler, Sarah Marler, Allyson Witters-Cundiff, Sanjeev Bhadresa, John Slattery, Juergen Hahn

## Abstract

**Purpose:** Several clinical studies have shown correlations between certain physiological measurements and an ASD diagnosis. Such findings, however, have generally not resulted in tangible progress towards practical translation due to a number of factors which this work seeks to address.

**Methods:** This paper presents a double-blind case/control trial design in which metabolic profiles, collected at two developmental pediatric clinics, were collected from children on a diagnostic waitlist for the purpose of developing a blood-based test for ASD. Besides obtaining blood samples, the children underwent gold-standard clinical evaluations, including the Autism Diagnostic Observation Schedule (ADOS), Mullens Scale of Early Learning (MSEL), and Vineland Adaptive Behavior Scale (VABS). The analysis, together with a complete medical history and physical exam, allowed to confirm or rule-out suspected ASD using DSM-5 criteria. The study was based on a cohort of 140 children between the ages 18-60 months, that were referred to a developmental pediatrician because of concerns in their development.

**Results:** 114 of these children received an ASD diagnosis, while 26 were diagnosed with non-ASD related developmental delays. Based on the measured metabolites, artificial intelligence-based classification algorithms allowed for an over 80% accuracy in predicting whether a sample came from a child diagnosed with ASD or not.

**Conclusion:** While these results need to be replicated in a larger study, especially involving more children with non-ASD related developmental delays, this is the first work using physiological measurements, coupled with AI, to support ASD diagnoses in a clinically relevant setting.

The clinical trial that was part of this work was registered on clinicaltrials.gov as NCT04672967 and was entitled the Metabolic Autism Prediction (MAP) Study. The study was IRB approved by the Biomedical Research Alliance of New York (BRANY) IRB on August 12, 2021 (approval number: A21-10-282-888).

## Introduction

Autism spectrum disorder (ASD) is a neurodevelopmental condition which is estimated to affect an ever-growing cohort of children in the United States, currently about 1 in 31 (3.2%) children [1]. In addition to the behavioral and health challenges for the individual diagnosed with ASD and their families, there is a significant socioeconomic burden with cost estimates ranging from $1M-$3M per individual [2] and $200B/year for national costs in the United States [3, 4]. In a separate study, the cost of ASD for the United States was estimated to be $3.6M per individual and a cumulative $4 trillion nationally from 2010 to 2019 [5].

Even though the American Academy of Pediatrics (AAP) recommends universal screening of ASD at 18- and 24-months well-baby visits [6], and given that many parents suspect a developmental problem at around 12 months of age, the average age of diagnosis remains at approximately 4 years [1, 7]. Moreover, at present ASD diagnoses are limited to observational, behavioral, and psychometric tools only. All diagnoses are made using the guidelines laid out in the Diagnostic and Statistical Manual of Mental Disorders, Fifth Edition (DSM-5) as provided by the American Psychiatric Association [8]. The gold-standard for ASD diagnosis is the Autism Diagnostic Observation Schedule (ADOS). The test must be administered by a trained clinician, often as part of a broader diagnostic workup with a developmental specialist.

This frequently results in a bottleneck for the ASD diagnostic process with long wait times to receive a diagnostic appointment. In the United States, the wait times are up to 24 months [9], and similar numbers are reported in other countries. Cortica (a clinical site involved in the current study being reported), for example, has a waitlist of 10 months.

Furthermore, disparities by race/ethnicity in estimated ASD prevalence and variations of the age of earliest comprehensive evaluation suggest access to treatment and services might be lacking or delayed for some children, creating healthcare disparities in accessibility for a timely diagnosis [10, 11]. The benefits of an early diagnosis, *i*.e., between 18-36 months, and an earlier behavioral intervention, however, are well documented. Behavioral intervention includes services, such as Applied Behavioral Analysis (ABA) or Early Intensive Behavioral Intervention (EIBI), which have demonstrated sustained improvements in IQ, adaptive behavior, challenging behavior, and generalized symptom severity when compared to children whose therapies started later [10, 12, 13].

Besides ASD symptoms being predominantly defined by difficulty in communication, social interaction, and restricted repetitive behaviors, individuals diagnosed with ASD also have a significantly higher likelihood to be diagnosed with other co-occurring conditions that affect a multitude of physiological systems [14, 15]. As the pathophysiology of ASD involves interactions between environmental and genetic factors, identifying distinctive physiological profiles of individuals with ASD has been a frequent subject of investigation [16]. An emerging way to evaluate ASD pathophysiology is through metabolomics, which can identify metabolic patterns of interest. These approaches generally fall into two broad categories: untargeted metabolomics [17, 18] and targeted metabolomics analysis [19, 20]. The former is characterized by measuring a large set of metabolites and identifying important differences in metabolite levels or patterns to discover disparities in metabolic pathways. Conversely, the latter approach postulates that certain pathways are predominantly affected and focuses on measuring metabolites of specific pathways. Targeted metabolomics is the approach used in this work.

This article puts emphasis on the folate-dependent one-carbon metabolism (FOCM) pathway, transsulfuration (TS) pathway, and energy metabolism. These pathways generate intermediate metabolites required for a wide variety of cellular mechanisms necessary for growth, development, and cellular homeostasis [21]. For example, these pathways influence gene expression via DNA methylation [22] and antioxidant/detoxification capacity via glutathione metabolism [23]. The reason for choosing these pathways is that DNA methylation has a direct effect on epigenetic expression which is deemed important for ASD [24], while glutathione plays a key role in oxidative stress which has also often been reported for children diagnosed with ASD [25]. More precisely, Mizejewski *et al*. found that biomarkers of oxidative stress in dried blood spots predicted an ASD diagnosis versus controls using immunoassays of newborn blood specimens [26]. Statistically significant differences in metabolite levels of these two pathways were first reported by James *et al*. [27]. In a further study by the same group, Melnyk *et al.* reported statistically significant differences in levels for several metabolites involved in FOCM and TS related pathways [23].

The data derived from one of their clinical studies [23] later used by Howsmon *et al*. to derive a machine learning based model that could predict whether the blood sample originated from a child with an ASD diagnosis or a typically developing child with an accuracy exceeding 96% [19]. A separate study, focusing on similar metabolites, verified these findings on a large cohort of children with ASD with over 88% accuracy [20]. In addition, the dataset from Melnyk *et al*. [23] was independently analyzed by Li *et al*. [28], confirming the results published in Howsmon *et al*.’s work [19]. Zou *et al*. performed a separate clinical trial involving metabolites of the FOCM/TS pathways and found that children with an ASD diagnosis could be separated from their typically developing peers with an accuracy exceeding 84% [29].

It is notable that in addition to FOCM/TS related pathways, additional measurements were used in this study. Specifically, alanine and lysine were added to the panel due to these analytes being surrogate biomarkers of impaired mitochondrial function which has been found in multiple ASD-related studies [30, 31, 32].

The main emphasis in this article is to address several challenges related to translating the promising academic findings related to abnormalities in FOCM/TS metabolites into a potential clinically validated commercial diagnostic test for predicting ASD in a clinical setting. This includes addressing the following challenges: (i) focus on distinguishing between children with an ASD diagnosis and those with developmental delays (DD), which is the commonly encountered case in a developmental pediatric setting, instead of the common practice in research to classify children with ASD and their typically developing peers, (ii) development of a new laboratory assay for measuring FOCM/TS metabolites using a commercially validated form of mass spectrometry to allow for accurate and precise quantification and commercial scale applications, and (iii) using clinical trial data collected prospectively at two independent trial locations pre-diagnosis, involving sample collection, preparation, and shipment, to simulate real world data collection. These are fundamental questions for developing a test for wider clinical use, which, thus far, has neither been developed nor discussed in the literature.

## Methods

This section describes the methods related to the cohort, measurements, and the analysis used for this study.

### Cohort Recruitment & Selection

The clinical trial that was part of this work was registered on clinicaltrials.gov as NCT04672967 and was entitled the Metabolic Autism Prediction (MAP) Study. The MAP Study was a double-blind case/control prospective clinical trial that enrolled 153 participants. Parents of participants provided informed consent. The study was IRB approved by the Biomedical Research Alliance of New York (BRANY) IRB on August 12, 2021 (approval number: A21-10-282-888). Participating sites included the Melmed Center (Cortica Care – Scottsdale location) and the Vanderbilt University Medical Center (VUMC). Both Melmed Center and VUMC offered children aged 18-60 months with a suspected developmental concern that were currently on their waitlist for ASD diagnosis an opportunity to take part in the study. Participants were considered eligible for the trial if they were primarily English speaking (due to the evaluation instruments being in English), parental consent (or legally authorized representative), if participants were willing to complete all study procedures and if the participants were not already enrolled in another clinical trial.

Children were ineligible for the trial if they had a previous diagnosis of ASD or developmental delay, premature < 34 weeks gestation, had a sibling enrolled in the clinical trial, had profound sensory deficits (*e*.*g*., hearing or vision deficits) that could interfere with interpretation of study results, documented or active seizures as defined by a clinical seizure or abnormal EEG within the past 6 months, one or more major genetic defects as determined by chromosomal microarray or other method of genetic detection, currently taking any high dose (greater than recommended daily allowance) dietary supplement, diagnosis or suspicion of viral/bacterial infection within 2 weeks of enrollment, diagnosis of congenital brain malformations, moderate to severe traumatic brain injury, congenital or acquired microcephaly, or infection of the central nervous system (e.g., meningitis or viral encephalitis), fetal alcohol syndrome, down syndrome, or other recognized syndrome usage of acetaminophen (Tylenol) within the past 72 hours, fever > 100 degrees Fahrenheit within the past 72 hours, or if the investigator felt participation in the trial may place the participant at an unnecessary risk.

The enrollment period for the study was from August 2021 through August 2023. All enrolled participants completed all study procedures, which consisted of a fasting blood draw and a full gold-standard diagnostic evaluation to rule-out an autism spectrum disorder. The diagnostic evaluation was performed by trained clinicians and consisted of the Autism Diagnostic Observation Scale, Vineland Adaptive Behavior Scale, Mullen Scales of Early Learning, modified Checklist for Autism in Toddlers, as well as a full medical history and physical exam to rule-out dysmorphology, and DSM-5 checklist to confirm ASD. The demographic information of the participants is summarized in Table 1 below. The intent of the trial was to ascertain whether metabolic profiles, collected in a double-blind manner, could predict idiopathic ASD for children that were awaiting a first-time gold-standard ASD diagnostic evaluation. Study physicians and evaluators were blind to metabolic results and laboratory personnel were blind to diagnostic information.

**Table 1.** Demographic Information of the Study Participants.

| Site | Cortica Care |  | Vanderbilt University Medical Center |  |
| --- | --- | --- | --- | --- |
| Clinical Category | ASD | DD | ASD | DD |
| # of Participants | 101 | 26 | 13 | 0 |
| Avg. Age (Months) | 40.8 ± 11.4 | 42.7 ± 11.9 | 37.4 ± 9.0 | N/A |
| Male (%) | 78.2 | 69.2 | 84.6 | N/A |
| Female (%) | 21.8 | 30.8 | 15.4 | N/A |
| Caucasian; not hispanic (%) | 34.7 | 42.3 | 46.1 | N/A |
| Caucasian; hispanic (%) | 28.7 | 26.9 | 23.1 | N/A |
| Asian American (%) | 9.9 | 7.7 | 7.7 | N/A |
| African American (%) | 7.9 | 0 | 7.7 | N/A |
| Other Races (%) | 18.8 | 23.1 | 7.7 | N/A |
| Race Not Specified (%) | 0 | 0 | 7.7 | N/A |

### Metabolites used for Classification

The folate-dependent one-carbon metabolism pathway encompasses a series of reactions which use folate (vitamin B9) to donate methyl groups. These folate-activated one-carbons are essential in several metabolic reactions, including the remethylation of homocysteine in the methionine cycle [33]. Within the methionine cycle, the methionine synthase reaction synthesizes methionine and tetrahydrofolate (THF), transferring a methyl group from 5-methyltetrahydrofolate (5-MTHF) to homocysteine [27]. Next in the cycle, methionine is activated into S-adenosylmethionine (SAM), which is involved in DNA methylation and acts as a methyl donor in various reactions. When SAM loses a methyl group, it becomes S-adenosylhomocysteine (SAH). SAH hydrolase (SAHH) then catalyzes a hydrolysis reaction, transforming SAH back into homocysteine and adenosine, which completes the methionine cycle.

Homocysteine can also be removed from the methionine cycle in an irreversible reaction that is catalyzed by cystathionine beta synthase (CBS), transforming it into cystathionine [27]. This reaction initiates the transsulfuration pathway. Cystathionine is then converted to cysteine in a reaction catalyzed by cystathionine gamma-lyase (CSE). Cysteine connects to glutathione synthesis, where the reduced form (GSH) and oxidized form (GSSG) of glutathione are in dynamic equilibrium [27, 34].

Previous studies have shown correlations of ASD with levels of these metabolites, reporting low ratios of SAM to SAH and GSH to GSSG [23]. People diagnosed with ASD have also shown increased levels of GSSG, as well as decreased levels of GSH, methionine, and cysteine [35]. There is also a correlation between ASD and oxidative stress, which is associated with DNA methylation, connecting back to metabolites in the FOCM pathway [25]. Similarly, abnormalities in folate metabolism and related metabolites can be related to ASD diagnosis [36]. Additionally, oxidative stress is known to be caused by mitochondrial dysfunction and oxidative stress itself can induce mitochondrial dysfunction [30, 37, 38]. Since impaired energy metabolism and mitochondrial dysfunction are important and connected pathways, levels of alanine and lysine were also evaluated to assess impairments of the energy metabolism. Additionally, studies have found decreased tryptophan metabolism [39] in children with ASD. Tryptophan plays an important role in the production of the neurotransmitter serotonin and in the regulation of the kynurenine pathway and its role in inflammation. Both serotonin and inflammation are known factors associated with ASD [40, 41, 42]. Tyrosine, which is a precursor for dopamine production and a metabolic byproduct of gut bacteria that can produce toxic compounds such as 4-ethylphenyl sulfate (4EPS) and its human analogue p-cresol, have been found to be at abnormal levels in children with ASD [43, 44, 45].

Seventeen metabolites and ratios of metabolites of the FOCM and TS pathways as well as TCA related amino acids were measured in this work. An illustration of the metabolic pathways and the metabolites can be found in Figure 1. The metabolites that are measured are highlighted in green. A detailed list of these measurements is given in Table 2 where the measurements are broken down into three groups (FOCM, TS, and TCA and/or Peripheral Neurotransmitter Precursor Measurements). All metabolites measured are used in the following analyses. Summary statistics are provided in Appendix Table 9. Normality testing is performed on the metabolite data using the Anderson-Darling test and skewness testing is performed using the D’Agostino’s test. The results can be found in the Appendix Table 10.

**Fig. 1.**
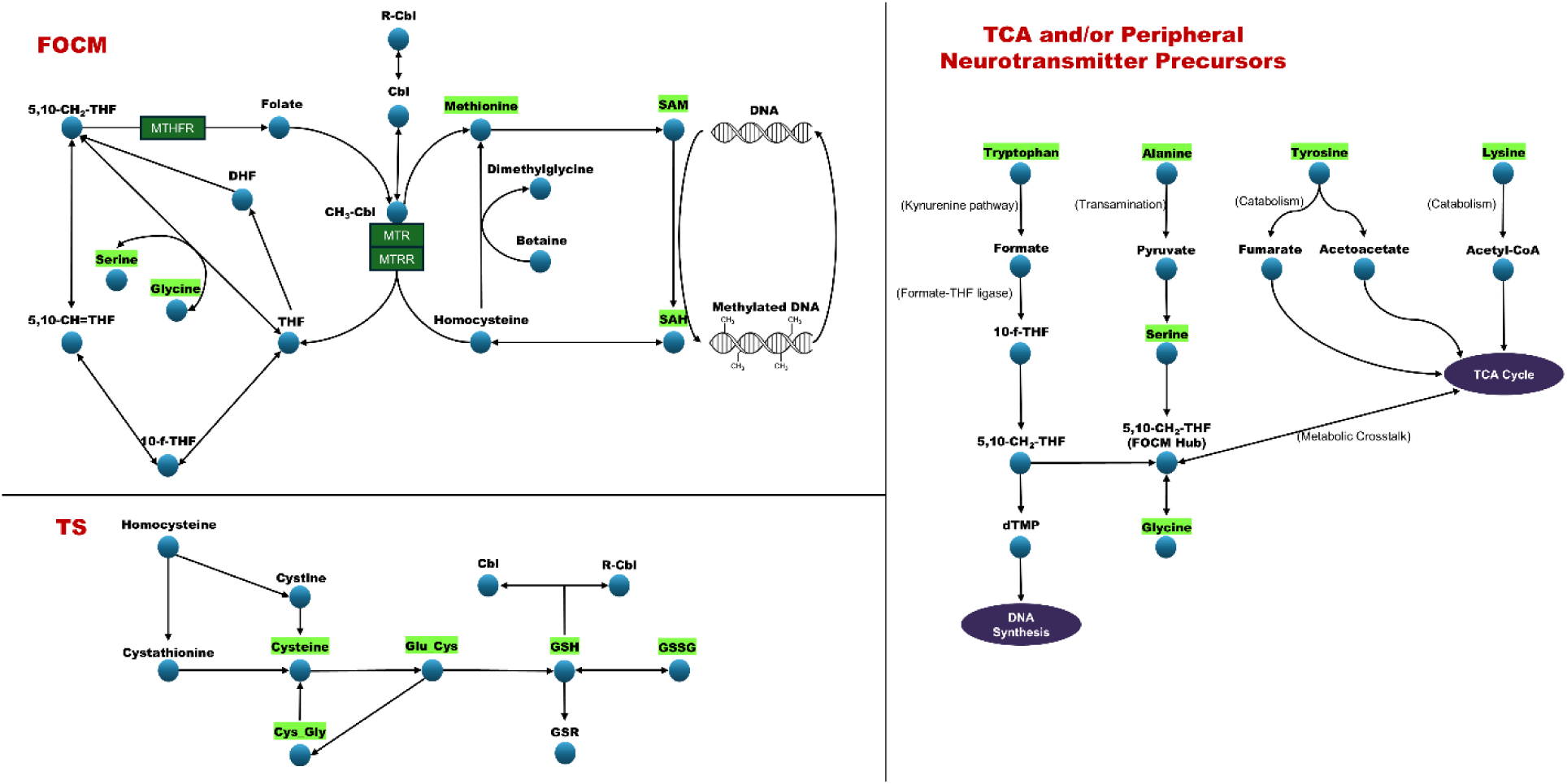
Associated pathways of metabolites measured in this study: FOCM (top left), TS (bottom left), and TCA and/or Peripheral Neurotransmitter Precursors (right). Metabolites in green indicate measurements taken in this work.

**Table 2.** Metabolites and Quantities Computed from Metabolites used in this Work.

| FOCM Measurements | TS Measurements | TCA and/or Peripheral Neurotransmitter Precursors |
| --- | --- | --- |
| Glycine | Cys_Gly_NEM (NEM stabilized L-cystenyl-L-glycine) | L-Alanine |
| L-Serine | L-Cysteine_NEM (free cysteine) | L-Lysine |
| L-Methionine | L-Cystine | L-Tryptophan |
| S-adenosylmethionine (SAM) | Glu_Cys_NEM (NEM stabilized (gamma-L-glutamyl-L-cysteine)) | L-Tyrosine |
| S-adenosylhomocysteine (SAH) | fGSH_NEM (free glutathione) |  |
| SAM/SAH | GSSG_T3 (oxidized form of glutathione) |  |
|  | %oxidized |  |

### Measurement Techniques

BioROSA Technologies, Inc. designed, developed, and validated a technological approach using LC-MS to interrogate analytes of interest. Since many of the analytes that have been shown to be abnormal in ASD are represented by a chemical class of molecules referred to as thiols, which are prone to oxidation and interconversion, BioROSA developed a stabilization approach using n-ethylmaleimide [46] to allow for accurate and precise quantification of all analytes, including thiols (*i*.*e*., GSH, cysteine, Cys-Gly, Glu-Cys all of which have a suffix _NEM to indicate derivatization). Instruments of choice included a SCIEX 5500 q Trap mass spectrometer and a Shimadzu LC-20 HPLC front-end. BioROSA completed an in-house technical validation prior to the beginning of the clinical trial. The results indicated that the assay was highly sensitive, specific, accurate, stable, and appropriate for clinical measurements. During the clinical study, the assay produced highly reproducible results for each patient sample, which were run maximum quadruplicate and a minimum of duplicate. All analytes had an average coefficient of variation (CV) of less than 15%, which is industry standard for analytical methods for clinical testing using LC-MS. Metabolic data was standardized across sites and sites used the same collection, storage, and shipping procedures. All metabolic data that were collected were run in batches with internal controls and run in triplicate to assess batch effects. There were no site related differences in data collected or run or site-specific differences in analyte concentrations.

### Data Preprocessing

The recorded dataset contained no missing data points and no data points were excluded. The processing steps involved classical standardization and dividing the set into training and validation sets to evaluate the generalizability of the trained classifiers. Eq. (1) shows how to standardize a recorded metabolite value, which is common practice [47]:

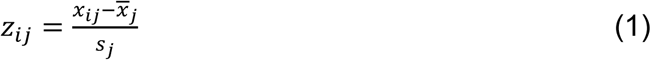

where *x_ij_* is the *i*th data point (measurement) of the *j*th metabolite, *x̅_j_* is the average (sample mean), *s_j_* is the sample standard deviation for all data points of the *j*th metabolite in the training set, and *z_ij_* is the standardized value of *x_ij_*, 1 ≤ *j* ≤ *m* and 1 ≤ *i* ≤ *N*_train_, with *m* = 17 and *N*_train_ being the number of metabolites used in this study and the number of data points (subjects) included in the training set, respectively. The standardized values have a sample mean of 0 and a sample standard deviation of 1 when computed on the training set. This technique was applied to the data before the feature engineering steps (see Section Feature Engineering) when the statistical features were calculated. No transformation, such as Log transformation, have been applied in this work.

### Linear Classification Models

A support vector machine (SVM) is a well-known binary classification method that determines an optimal hyperplane (dividing plane) to separate two classes of data points. Based on the widest margin approach, the SVM algorithm computes the optimal hyperplane, ℎ(*z*), as a linear combination of the metabolites and an offset (bias) term, as shown in Eq. (2):

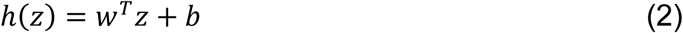

where *z* is a standardized *m*-dimensional data vector storing the recorded values of the *m* metabolites, *w* denotes a weight vector that determines the orientation of the dividing hyperplane and the bias term *b* influences the distance of the hyperplane from the origin, in the data or feature space [48]. For linear kernels (or scalar products of data points 1 ≤ *i*, *j* ≤ *N_train_*), which Eq. (3) shows, the dividing hyperplane is designed for the original data space.

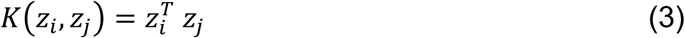

The results reported in this work were based on the SVM approach, described above, as a benchmark. A nonlinear extension of SVM is introduced below.

### Nonlinear Classification Models

An extension of the linear SVM approach is based on a nonlinear transformation of the original (standardized) metabolites, which is of the form:

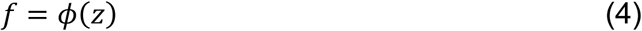

Here, *f* is the nonlinearly transformed variable (metabolite) set *z*, and *φ*(·) is a nonlinear function to generate the feature vector *f*. Put differently, based on the Cover theorem [49], nonlinearly transforming the original data into a high dimensional feature space improves the separability for nonlinearly separable classes. By virtue of its construction, the linear kernel function introduced in Eq. (3) can easily be augmented by replacing the scaled data vector with its nonlinearly transformed version, which gives rise to:

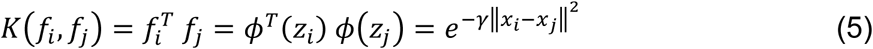

This nonlinear version is often referred to as kernel SVM or kSVM [50]. Other than changing the kernel function from the linear kernel to a nonlinear kernel in Eq. (5), kSVM is based on the same algorithm as the linear SVM one. Eq. (5) highlights that the scalar products are expressed by an exponential function (Gaussian kernel) that has a squared and scaled argument with *γ* being the scaling factor. The performance of a kSVM classifier depends on the selection of the kernel parameter [50] which was performed via an exhaustive grid search in this work.

### Feature Engineering

Given the small number of participants with a developmental delay diagnosis in the clinical trial, this dataset is, consequently, unbalanced, rendering the classification task challenging. To address this, a feature engineering approach was applied [51]. In short, this approach relies on manually computing engineered features which are derived from the existing measurements and, subsequently, apply kSVM based on the engineered features.

The original 17 metabolites can be divided into the following four groups: (i) FOCM, (ii) TS, (iii) TCA and/or Peripheral Neurotransmitter Precursors, and (iv) “All”. FOCM and TS include the metabolites from those specific pathways, and TCA and/or Peripheral Neurotransmitter Precursors includes selected metabolites related to the energy metabolism, as listed in Table 2. The “All” category includes all 17 metabolites. The dataset contains two already-computed features: SAM/SAH in the FOCM pathway and %oxidized in the TS pathway. These calculated metabolites are included in their respective groups. Within these four groups of metabolites, classical summary statistics were computed: (i) arithmetic mean, (ii) arithmetic standard deviation, (iii) geometric mean, (iv) geometric standard deviation, (v) entropy, (vi) sum, (vii) variance, (viii) skewness, (ix) kurtosis, (x) median, quantiles (xi) 5 and (xii) 95, (xii) CV (coefficient of variation), (xiv) range, (xv) IQR (interquartile range), and (xvi) energy, which represent the features used in this study. These 16 statistics were calculated, per participant, for each of the four groups, resulting in a total of 64 features.

Based on the FOCM and TS pathways, as shown in Figure 1, a number of metabolites are used to derive additional features. When independently investigating single metabolites, small alterations may seem physiologically irrelevant, but they can impact the function of related enzymes and transporters [18]. Creating ratios between multiple metabolites involved in reactions can be an indicator of changes in reaction activity in a pathway. Such ratios can also normalize metabolic measurements and reduce systemic experimental errors [52]. A similar technique was used by Melnyk *et al*., where ratios of SAM to SAH and GSH to GSSG were investigated and they reported statistically significant differences in the ratios among different groups [23].

This type of technique is employed here to analyze the biochemical relationships between different metabolites that appear in Figure 1, creating several additional features which are described in Table 6. This resulted in an additional 42 features for a total of 123 features, including the initial 17 features, 64 statistical features, and the 42 biochemical relation features, all of which can be used as potential inputs for a classifier. A full list of the original features and their respective metabolic pathways is shown in Table 2. The engineered features added to the dataset are listed in Tables 7 and 8 in the supplemental materials section.

### Leave-One-Out Cross-Validation Approach

This validation approach is based on a holdout strategy, where the *N* data points are systematically divided into a training set of *N* − 1 data points and a validation set containing the single data point that is not included in the training set [53]. This approach gives rise to *N* trained models and commences by leaving the first data point out, then the second one and so on. The data point left out is used to validate the performance of the model trained on the *N* − 1 data points of the training set. Leave-One-Out Cross-Validation (LOOCV) is a potential remedy for evaluating the generalizability of trained models when only a small number of data points are available. It is, however, advisable to consider additional model evaluation, as LOOCV does limit the validation set to a single data point only. This single data point limitation may not allow for evaluating the generalizability of a linear or nonlinear model This study relies on applying LOOCV to evaluate the generalizability of kSVM classifiers and an additional Monte Carlo evaluation, which is described next.

### Monte Carlo Simulations

To introduce a more rigorous assessment into the ability of separating the two classes (ASD vs DD), this study also utilizes Monte Carlo simulations, which are based on repeated random sampling [54]. More precisely, instead of utilizing a single data point, the Monte Carlo concept relies on random train/test splits of the data. This study considered 80/20 train/test splits for most of this study. In addition to that, train/test splits of 85/15 and 90/10 were also investigated to explore the effects of including more data points in the training set. The number of repetitions (selecting random splits) was set at 5,000 to address the small sample size and the imbalance between the ASD and DD classes. For each Monte Carlo run, the performance metrics of the model are computed to evaluate its predictive capability of that model. The mean and standard deviation of each performance metric across all simulations are computed and reported.

### Feature Selection

This work used an exhaustive search to evaluate the performance of combinations of four features for each trained classification model. When extending to five features, however, evaluating all combinations become too computationally expensive, *i*.*e*., the number of combinations of five out of 123 features is 216,071,394. In comparison to that, an exhaustive search for four features required 9,078,630 combinations which already required approximately 5.5 hours on an Intel Core i7-7800X CPU when running Python 3.10.11.

Instead, a greedy approach was employed for constructing classification models for subsets of five features. A greedy approach is based on a predetermined set of features to which one of the not already selected features is added [55]. This study applies the greedy approach by taking the 4-feature kSVM classification model with the highest balanced accuracy and testing the addition of the remaining 119 features in a one-at-a-time mode. Each of these models, containing 5 features now, was evaluated using both LOOCV and Monte Carlo simulations.

An exhaustive search of all feature combinations of up to four features was used with a kSVM model. The top 10,000 feature combinations, per train balanced accuracy, of this search were re-evaluated using a LOOCV approach to evaluate the model performances in a deterministic manner, and the results were ranked by the balanced accuracy of the test set. The four feature model with the highest balanced accuracy was used in the greedy approach as explained above. All data preprocessing steps, feature selection via exhaustive and greedy search and hyperparameter tuning are performed prior to cross valudation.

To evaluate the classification models, this study relied on balanced accuracy, the area under the receiver operator characteristic (ROC) curve, accuracy, sensitivity, specificity, precision, and the F1 and MCC scores. The results section lists the values of these metrics for the best performing models.

## Results

The analyzed dataset is comprised of data from 140 individuals: 114 individuals with an autism spectrum disorder (ASD) diagnosis and 26 individuals diagnosed to be developmentally delayed (DD). As a baseline, linear SVM classifiers were trained first, based on the original 17 metabolite features. An exhaustive feature search to include up to 5 features (out of 17) was performed to determine the inputs to the SVM model. The results confirm that linear classification models failed to accurately distinguish between the two classes, as the best performing linear classification model, evaluated using the LOOCV method, attained a balanced accuracy of 50%, an accuracy of 81.43%, a sensitivity of 100%, and a specificity of 0%. For sake of completeness, other linear classification models, such as Fisher Discriminant Analysis and logistic regression were also applied and returned the same results. These results are supported by scatterplots of each feature, shown in Figure 6 in the Appendix. The values of each data point are relatively scattered, and no clear difference is noticeable between the ASD and DD cohorts, indicating that the two groups are not linearly separable.

The next step was training nonlinear kSVM classifiers based on the 106 engineered features, detailed in Section Feature Engineering, and the original 17 metabolites. As highlighted in Section Feature Selection, the resultant exhaustive combinatorial search attained the best sets of four feature combination and the greedy search approach obtained the best performing five feature sets, based on the balanced accuracy. As discussed in Sections Leave-One-Out Cross-Validation Approach and Monte Carlo Simulations, the performance of each model was assessed using LOOCV and the Monte Carlo scheme. Tables 3 and 4 detail the performance of the most accurate classification models (average for the Monte Carlo runs is used here). It is important to note that, although Tables 3 and 4 list the metrics of the best performing models, a similarly high balanced accuracy was obtained by numerous combinations of the 4 or 5 features of the original set of 123.

**Table 3.** Results of 4-feature Models with LOOCV and Monte Carlo (mean and standard deviation)

| Method | Bal-<br>anced<br>Accu-<br>racy | AUC | Accu-<br>racy | Sensi-<br>tivity | Specific-<br>ity | Preci-<br>sion | F1 | MCC |
| --- | --- | --- | --- | --- | --- | --- | --- | --- |
| LOOCV | 0.8644 | 0.8853 | 0.9000 | 0.9211 | 0.8077 | 0.9546 | 0.9375 | 0.6907 |
| Monte<br>Carlo | 0.7819<br>±<br>0.1093 | 0.8602<br>±<br>0.0918 | 0.8672<br>±<br>0.0580 | 0.9145<br>±<br>0.0560 | 0.6493<br>±<br>0.2150 | 0.9250<br>±<br>0.0433 | 0.9184<br>±<br>0.0365 | 0.5615<br>±<br>0.2005 |

**Table 4.** Results of 5-feature Models with LOOCV and Monte Carlo.

| Method | Bal-<br>anced | AUC | Accu-<br>racy | Sensi-<br>tivity | Speci-<br>ficity | Preci-<br>sion | F1 | MCC |
| --- | --- | --- | --- | --- | --- | --- | --- | --- |
|  | Accu-<br>racy |  |  |  |  |  |  |  |
| LOOCV | 0.8731 | 0.9001 | 0.9143 | 0.9386 | 0.8077 | 0.9554 | 0.9469 | 0.7255 |
| Monte<br>Carlo | 0.8007<br>$\pm 0.1083$ | 0.8750<br>$\pm 0.0918$ | 0.8837<br>$\pm 0.0544$ | 0.9298<br>$\pm 0.0500$ | 0.6715<br>$\pm 0.2137$ | 0.9306<br>$\pm 0.0428$ | 0.9291<br>$\pm 0.0336$ | 0.6088<br>$\pm 0.1937$ |

Table 3 shows the results of evaluating the best 4-feature combination, per balanced accuracy, computed using LOOCV and the average of the 5,000 Monte Carlo runs for an 80/20 train/test split. The LOOCV model yielded a balanced accuracy of 0.8644, and the Monte Carlo evaluation a balanced accuracy of 0.7819. As expected, the LOOCV evaluation strategy attained a higher balanced accuracy as compared to the Monte Carlo approach, given that LOOCV relies on a smaller test set (only a single data point) and a larger training compared to the Monte Carlo scheme. The AUC value of model followed the same pattern. The AUC values are 0.8853 and 0.8602 when using the LOOCV and Monte Carlo evaluation approaches, respectively. The accuracy of the models when evaluated based on the LOOCV and Monte Carlo schemes, respectively, were 0.9 and 0.8672, and the MCC values were 0.691 and 0.562, respectively. The confusion matrix of the 4 feature LOOCV model is shown in Figure 2.

**Fig. 2.**
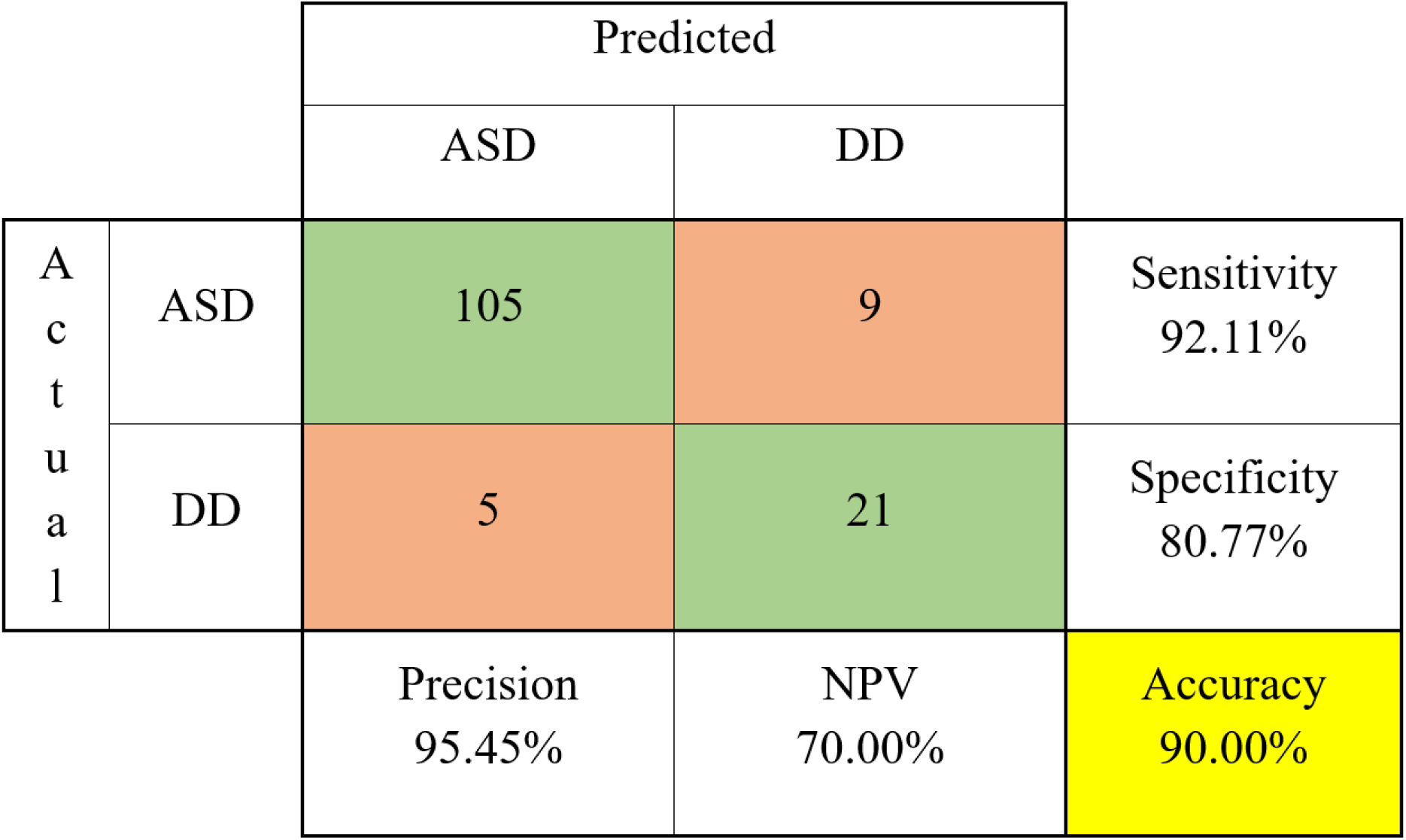
Confusion Matrix of 4-feature Model with LOOCV (NPV – negative prediction value, all numerical values in percentage instead of fractions)

To support the AUC value of 0.89 for the 4-feature LOOCV model presented in Table 3, the ROC curve of this model is displayed in Figure 3(a). The curve shows significant increases in the true positive rate at low false positive rate values, indicating a good model fit. The ROC curve for the 4-feature combination with 5000 Monte Carlo simulations is shown in Figure 3(b), with an AUC value of 0.86. There is still a significant rise in the true positive rate at lower values of the false positive rate. As there are only 5 DD values in the test set per Monte Carlo run with 20% testing set size, there are only 6 possible values the false positive rate can take. The feature set with the highest balanced accuracy is the same for both LOOCV and Monte Carlo simulations, which is ts_skewness, SAM_div_sum_Methionine_SAH, focm_mean, SAM_minus_SAH. The details about the features can be found in the supplemental material.

**Fig. 3.**
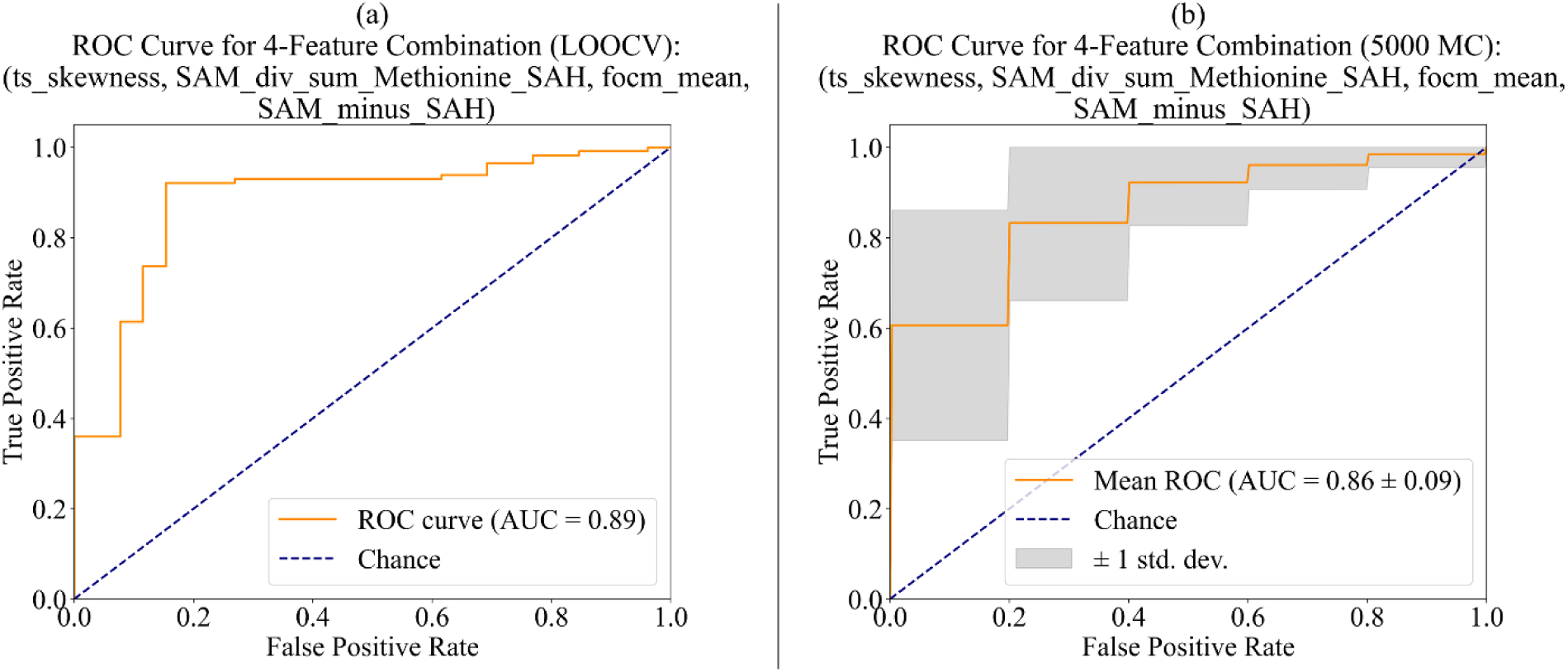
ROC Curve of 4-Feature kSVM Classifier: (a) LOOCV, (b) average over 5000 Monte Carlo runs.

The results of applying the most accurate 5-feature combination models, per balanced accuracy, with LOOCV and Monte Carlo simulations with an 80/20 train/test split, are shown in Table 4. The model evaluation using LOOCV produced a balanced accuracy of 0.8731 and that of a Monte Carlo evaluation yielded an average of 0.8007. The models attained an area under the receiver operator characteristic curve (AUC) of 0.9001 and 0.875 based on applying a LOOCV and Monte Carlo-based assessment, respectively. In addition to that, an accuracy of 0.9143 and 0.8837 can be reported for the models for an evaluation using LOOCV and the Monte Carlo method, respectively. The difference in AUC value and accuracy is approximately 3% different between the two models. The MCC values are 0.7255 and 0.6088 for an LOOCV and a Monte Carlo evaluation. Figure 4 below presents the confusion matrix for the 5-feature model with LOOCV.

**Fig. 4.**
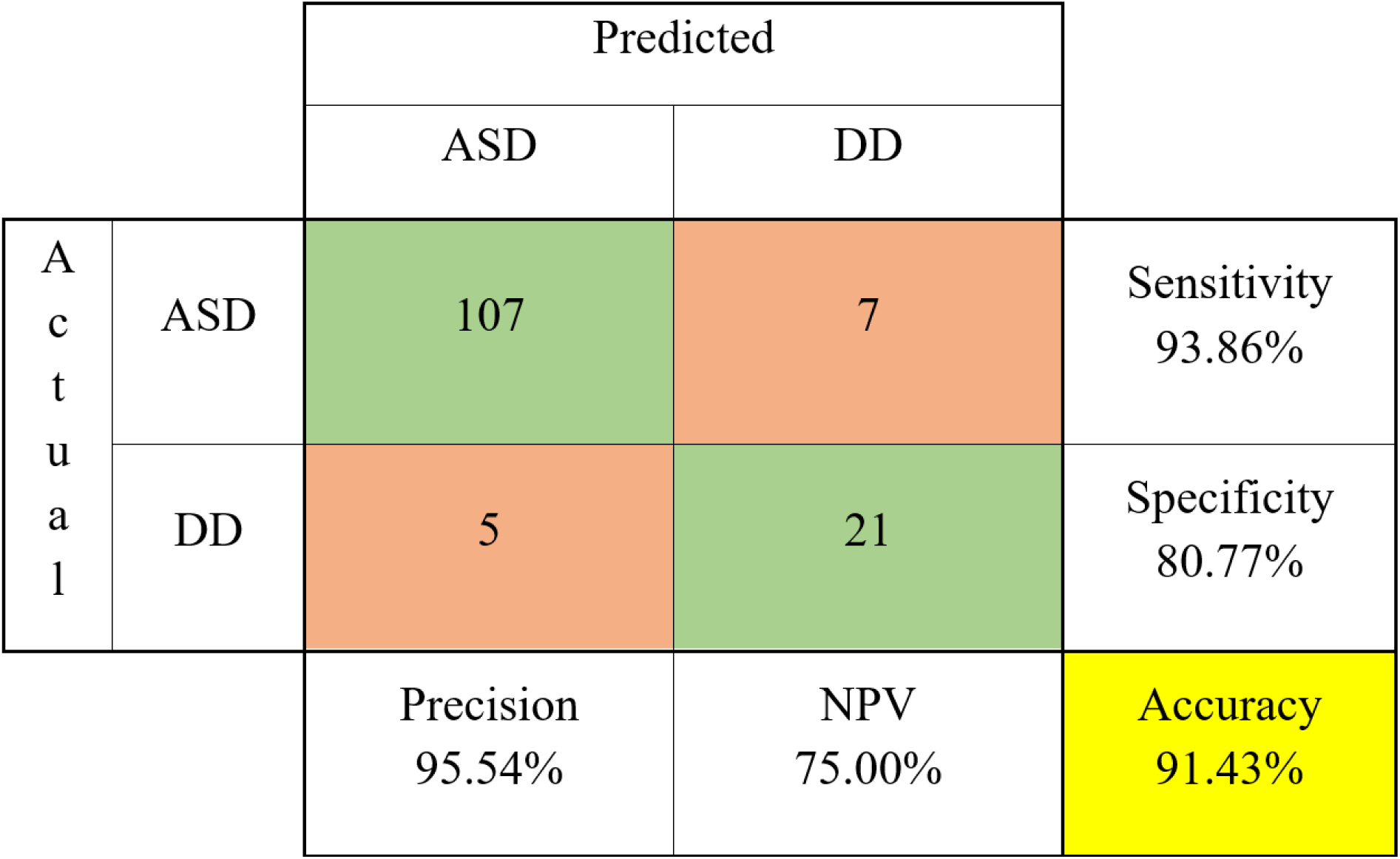
Confusion Matrix of 5-feature Model with LOOCV.

The receiver operator characteristic (ROC) curve of the 5-feature model (LOOCV evaluation) is shown in Figure 5(a). This curve also has a significant spike in the true positive rate at low false positive rate values, indicating good model performance. The receiver operator characteristic curve of the 5-feature model (5000 run Monte Carlo evaluation) is shown in Figure 5(b). Figure 5(b) also highlights considerable increases in the true positive rate at low false positive rate values. The additional feature of the 5 feature models, determined by a greedy search, was avg_SAM_SAH_SAMSAH (Table 6, Eq. A.3) and was the same for both LOOCV and Monte Carlo simulations.

**Fig. 5.**
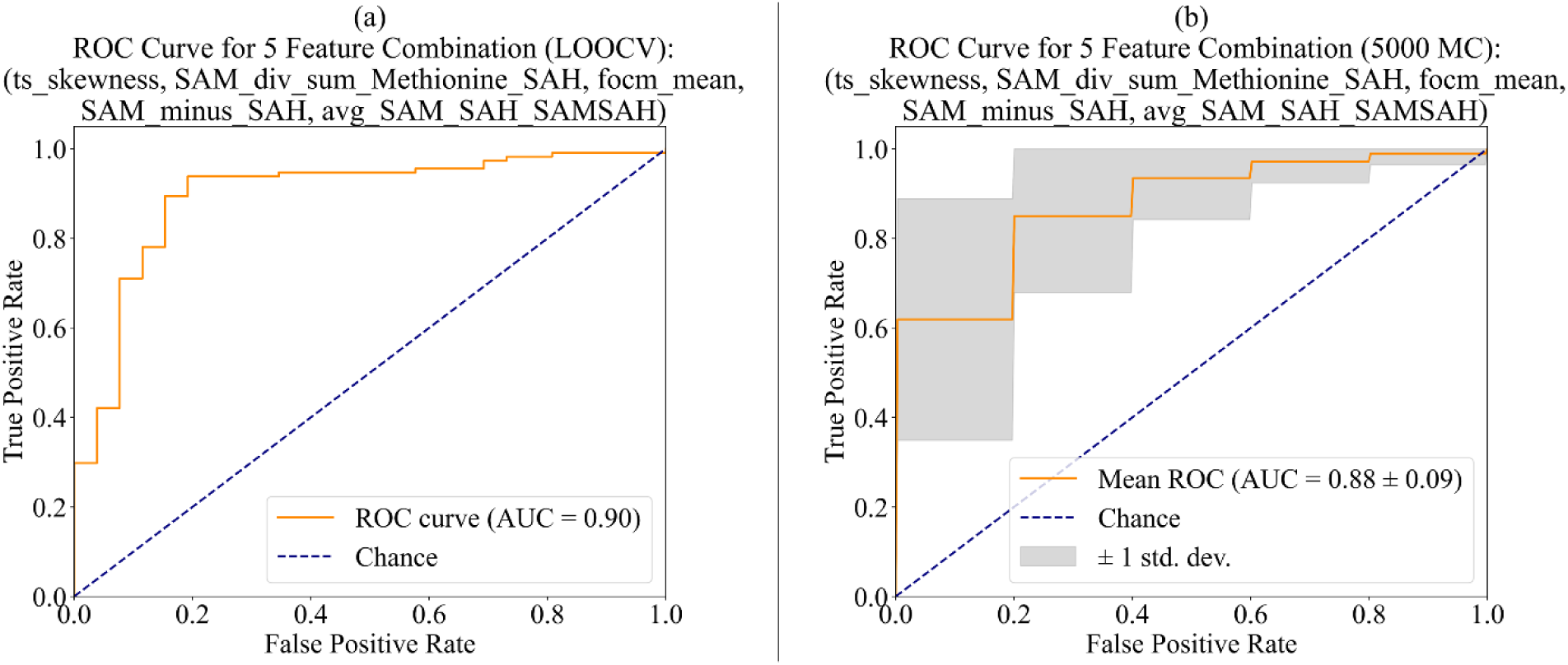
ROC Curve of 5-Feature kSVM Classifier: (a) LOOCV, (b) average over 5000 Monte Carlo runs.

Comparing the results of the 4-feature and 5-feature models, the LOOCV had balanced accuracy and accuracy values of approximately 0.86 and 0.9 for the 4-feature model, and approximately 0.87 and 0.91 for the 5-feature model. This minor but noticeable difference in each performance metric was also seen in the Monte Carlo simulations when comparing the 4- and 5-feature models, suggesting that adding the fifth feature marginally increased the accuracy of the model and that including additional features may not result in a considerable increase in each performance metric.

Overall, the high classification accuracy attained by both the 4- and 5-feature models, compared to the linear baseline models confirms that separation between the two groups of ASD and DD individuals, based on metabolites from the FOCM and TS pathways as well as selected energy metabolism related metabolites, is possible. In other words, these metabolites support discerning an ASD from a DD pre-diagnosis in a clinical setting for children showing signs of developmentally impairment.

Another important aspect for this study is the imbalance in the dataset. To address this, the effect of different train/test splits upon the results of the Monte Carlo evaluation was investigated for 80/20, 85/15, and 90/10 train/test splits, with a particular focus on specificity. Table 5 shows the average specificity values, which tendentially increased with larger training sets/smaller test sets. This suggests a stronger predictive power for DD individuals as more of them were included in the training set. This observation confirms that significant dependence upon the small number of DD samples, which constitutes a limiting factor for this study. However, it is also important to note that, despite this imbalanced dataset, a high classification accuracy (balance accuracy) can be attained, which also confirms the validity and merit of developing clinical tests for a pre-diagnosis in young children with behavioral issues.

**Table 5.** Specificity Values of Monte Carlo Simulations with varying Train/Test Splits.

| Number of Features | Method and train/test split | Specificity |
| --- | --- | --- |
| 4 | Monte Carlo with 80/20 | 0.6493 |
|  | Monte Carlo with 85/15 | 0.6748 |
|  | Monte Carlo with 90/10 | 0.6971 |
| 5 | Monte Carlo with 80/20 | 0.6715 |
|  | Monte Carlo with 85/15 | 0.6928 |
|  | Monte Carlo with 90/10 | 0.7093 |

## Discussion

In this study, targeted metabolomic analysis is used to evaluate classification of children with autism spectrum disorder and children with other non-ASD related developmental delays, based upon their metabolic profiles. It was possible to build classification models with high diagnostic accuracies based on metabolites from the folate-dependent-one-carbon metabolism, transsulfuration pathway, and energy metabolism which are measured through mass spectrometry. The top four feature kSVM model achieved a LOOCV balanced accuracy of 86.4%, accuracy of 90.0%, AUC of 0.885, and MCC of 0.691. Across 5000 Monte Carlo simulations with an 80/20 train/test split, a balanced accuracy of 78.2%, accuracy of 86.7%, AUC of 0.860, and MCC of 0.562 was achieved. Incorporating a fifth feature yielded modest improvements with a LOOCV balanced accuracy of 87.3%, accuracy of 91.4%, AUC of 0.900, and MCC of 0.726. With 5000 Monte Carlo simulations, the 5-feature model reached a balanced accuracy of 80.0%, accuracy of 88.3%, AUC of 0.875 and MCC of 0.609. These results show that it is possible to distinguish between children with an ASD diagnosis and those with a non-ASD related developmental delay based upon measuring a limited number of metabolites through a scalable mass spectrometry technique. The results also indicate that four features were able to capture the majority of the difference between the two classes and adding a fifth feature modestly improved the results.

Howsmon *et al*. has achieved over 96% accuracy using linear Fisher’s discriminant analysis to differentiate ASD from typically developing individuals [19]. However, in this work, linear approaches have failed to result in feasible classifiers as the investigated scenario is a more challenging task. This is likely the case because the metabolic profiles of the DD cohort also involve dysregulation of the pathways of interest, *i*.*e*., this DD group introduces more unknowns and challenges into the classification task as opposed to the TD metabolic profiles. This work also uses a measurement design where samples are taken at multiple sites and measurements are processed at another site, which induces more logistical variation but simulates more realistic multi-site procedure, unlike the data in Howsmon *et al.* [19, 23]. As can be seen in the scatter plots of the data in Figure 6 in the appendix, there is neither a separation between nor a clustering of the data points among the ASD and DD cohorts in any of the measurements, unlike the data in the Howsmon *et al.* where there were clear separations between the ASD and TD cohorts in many of the measurements even though there is some overlap [19]. The fact that there is no statistical difference between ASD and DD cohorts makes it infeasible to use linear classification models. However, applying Cover’s theorem allows for discovering complex patterns that render the classification problem is more likely to be linearly separable when mapped into a higher-dimensional space [49]. Thus, more sophisticated methods were needed to achieve the accuracies reported in this work.

Similarly, nonlinear classification approaches, based on the original 17 metabolites, have struggled to appropriately learn the relationship between ASD and DD cohorts since the number of DD points is low at only 26 samples. kSVM implicitly maps the data into a higher dimensional space where a linear separation is possible. The model needs to infer the separation structure from the distances among the data points. Designing feature engineering before applying kSVM allows the model to learn the separation structure from less data as some structure is already encoded in the input, so the kSVM model is trained on such predefined and useful pattern solely from pairwise distances. Thus, the kSVM can learn to construct a separating boundary with few samples.

The reported results extend earlier targeted metabolomic studies based on FOCM and TS pathways [19, 20, 28, 29], which reported 84-96% accuracy for distinguishing children with ASD from typically developing peers. The reason for difference in accuracies in this work is that the task of differentiating ASD vs DD is more challenging than ASD vs TD, given that the developmental delays are not well characterized and may include some physiological commonalities with ASD at the biological pathway level. It is important to point out that the classification of ASD vs developmental delay is of high clinical relevance as children evaluated for ASD are generally seen by a specialist precisely because there are concerns regarding their development. As such, the often investigated classification of ASD vs typically-developing children is of limited applicability in a clinical setting for early diagnosis and early intervention. Children that are referred to a diagnostic center for suspected ASD, by definition, have suspicions of developmental concerns, which are also, by definition, not present in children who show a typical development.

The high values of the performance metrics reported in this study underscore the potential of the FOCM/TS pathway-derived metabolites, in combination with engineered features, to capture the difference between children with ASD from those with DD. This aligns with other studies that show that dysregulation in FOCM and TS pathways is correlated with ASD pathophysiology [56]. The findings reported here support that perturbations in methylation capacity and glutathione-dependent redox homeostasis are commonly found in ASD [19, 27, 57, 58] and demonstrate that these disturbances remain detectable against the metabolic background of non-ASD developmental delays.

From a translation perspective, this work focused on three key challenges. First, the use of a scalable mass-spectrometry laboratory protocol supports potential clinical deployment [59]. Second, the focus on distinguishing ASD from DD individuals instead of ASD vs TD brings the context of the research to align more closely with real-world clinical scenarios. Lastly, the study design with sample collection at multiple sites and measurement processing at another site demonstrates resilience to logistical variations that could impede multi-center implementation. These advancements could move the field closer to a practical blood-based supplementary test for ASD screening and diagnosis in developmental pediatric clinics. Another strength of this work is the prospective double-blind case/control design. While this was important to minimize bias and create a strong clinical trial design, it also created a significant imbalance between ASD cases and DD controls. Future work will need to consider more balanced datasets while also maintaining a rigorous clinical trial methodology.

To put these findings into the context of developments of physiological tests for ASD, Quadrant Biosciences has commercialized a test based on salivary miRNA that aims to diagnose ASD [60, 61, 62]. In their 2016 study, the salivary miRNA was purified from 24 ASD and 21 TD individuals. The top fourteen miRNAs were used to fit classification models which achieved AUC values over 0.91. In the validation study published in 2018, 456 children in total from multiple centers were recruited of which 134 were TD, 238 were ASD, and 84 with non-ASD developmental delays. This 2018 study achieved an accuracy of 85% based on thirty-two salivary RNA features while classifying ASD vs non-ASD. In the follow up study published in 2019, twenty-eight salivary miRNAs were identified. The classification algorithm using four miRNAs to differentiate ASD vs non-ASD achieved 89% sensitivity and 32% specificity. While these results look promising, the test offered by Quadrant Biosciences was discontinued during the COVID-19 pandemic. Furthermore, NeuroPointDx offers a blood test that measures blood metabolites and compares the levels of the individual to the signature metabolic profiles of children with ASD in their clinical study named Children’s Autism Metabolome Project [32]. Their test provides information about the metabolism of the child and recommends further testing if the individual has metabolic signature associated with ASD; however, it is not a standalone diagnostic test. Additionally, the work by Flynn et al. found that some microbiome produced metabolites have unusually high levels in the urine of children diagnosed with ASD [63]. While this is not an approved test for ASD, the assay that this work is based on is commercially available in the UK [63]. No other test, currently available, to diagnose ASD exists at this time.

Like any study, there are several limitations for this work. First, even though the ASD cohort was reasonably large with 114 individuals, the DD cohort was relatively small with 26 individuals, resulting from a high rate of the recruited children being diagnosed with ASD. This created an imbalance that made it more challenging for classification models to identify an optimal separation boundary. This is also highlighted by the fact that the specificity values increased from 65% to 70% with increasing training split sizes in the Monte Carlo simulations. Future work should prioritize expanding the DD cohort to minimize the class imbalance and allow for a richer data set of individuals in the DD category. Additionally, data collection locations can be diversified to better capture any bias or variation resulting from factors varying from one clinical site to another. Furthermore, the possibility of overfitting due to the addition of feature engineering step cannot be completely ruled out, even though the consistency of performance between LOOCV and Monte Carlo simulations shows the models’ robustness across training-set variability. While the engineered statistical features selected by our model demonstrated robust stability during cross-validation, their nature limits direct biological interpretability. Because they are mathematically derived rather than direct biomarkers, future studies utilizing independent, external cohorts are necessary to confirm their broader generalizability. Finally, future work should explore whether an even smaller raw-metabolite panel could match the performance, especially once class imbalance is addressed through collection of additional DD data points.

Another limitation is that the Vanderbilt study site yielded only individuals diagnosed with ASD, with no cases of developmental delay. Furthermore, the sample size from this site was limited to just 13 individuals, which may raise concerns regarding potential site-specific effects. To address this, we conducted a supplementary analysis excluding these 13 data points, utilizing solely the data from Cortica Care (101 ASD and 26 DD cases). No significant changes were observed when leaving out the data from the smaller of the two sites.

Overall, this work is the first and only paper that highlights that targeted metabolomics of the FOCM/TS pathways and energy metabolism, coupled with an analysis based upon data science, can distinguish between ASD and other developmental delays with accuracies exceeding 80%. While these findings represent significant contributions toward translation, establishing true readiness for clinical deployment will still require future external validation, reproducibility across centers and labs, calibration analysis, and prospective utility testing.

This prospective double-blind study has established a clinically meaningful framework for blood-based ASD biomarker research. By recruiting children from diagnostic waitlists at two geographically distinct pediatric clinics and performing gold-standard blinded clinical evaluation, the study minimizes retrospective bias and maximizes clinical relevance to first-time diagnostic evaluations. It has demonstrated that a panel of FOCM/TS and energy metabolism metabolites, which were augmented by feature engineering, can distinguish children with ASD from those with other developmental delays with balanced accuracies exceeding 80%. Through the implementation of a scalable mass-spectrometry workflow, domain-driven feature construction with kSVM, and validation across geographically separated collection and measurement sites, this work has addressed several critical barriers to clinical deployment of blood-based ASD support diagnostics. These barriers include the limited clinical relevance of ASD vs TD discrimination in previous works, the lack of scalability of laboratory-specific measurement protocols, the retrospective nature of studies, and the challenge of robust implementation across multiple collection and analysis sites. The consistency between LOOCV and extensive Monte Carlo simulations reveals model robustness to training-set variability. The limited sample size and imbalance between the ASD and DD cohorts impeded classification and posed challenges for standard linear and nonlinear classification techniques. However, the combination of kSVM with feature engineering was able to deal with the limited number of samples from non-ASD related developmentally delayed children and achieve good classification accuracy. These results provide a step towards the development of a scalable blood-based support diagnostic test for early ASD screening.

## Data Availability

All data produced in the present study are available upon reasonable request to the authors

## Statements and Declarations

### Ethics Approval

The clinical trial that was part of this work was registered on clinicaltrials.gov as NCT04672967 and was entitled the Metabolic Autism Prediction (MAP) Study. The study was IRB approved by the Biomedical Research Alliance of New York (BRANY) IRB on August 12, 2021 (approval number: A21-10-282-888). The procedures used in this study adhere to the tenets of the Declaration of Helsinki.

### Consent

Informed consent was obtained from all participants as part of the trial.

### Data

Data is stored at Rensselaer Polytechnic Institute, Troy, NY. Data will be made available upon request.

### Contributions

**Halil Arici** (Conceptualization, Methodology, Software, Validation, Formal Analysis, Investigation, Data Curation, Writing – Original Draft, Writing – Review & Editing, Visualization, Supervision, Project Administration), **Marie Causey** (Conceptualization, Methodology, Investigation, Resources, Data Curation), **Soma Patra** (Conceptualization, Methodology, Software, Validation, Formal Analysis, Investigation, Data Curation, Writing – Original Draft, Writing – Review & Editing, Visualization), **Uwe Kruger** (Conceptualization, Methodology, Investigation, Resources, Data Curation, Writing – Original Draft, Writing – Review & Editing), **Cristopher Antonio Villegas Uribe** (Software, Formal Analysis), **Raun Melmed** (Investigation, Resources, Data Curation), **Craig Ciuk** (Investigation, Resources, Data Curation), **Sophia Crisler** (Investigation, Resources, Data Curation), **Sarah Marler** (Investigation, Resources, Data Curation), **Allyson Witters-Cundiff** (Investigation, Resources, Data Curation), **Sanjeev Bhadresa** (Conceptualization, Methodology, Investigation, Resources, Data Curation), **John Slattery** (Conceptualization, Methodology, Investigation, Resources, Data Curation, Writing – Original Draft, Writing – Review & Editing, Supervision, Project Administration, Funding Acquisition), and **Juergen Hahn** (Conceptualization, Methodology, Investigation, Resources, Data Curation, Writing – Original Draft, Writing – Review & Editing, Supervision, Project Administration, Funding Acquisition)

## Appendix

**Fig. 6(a).**
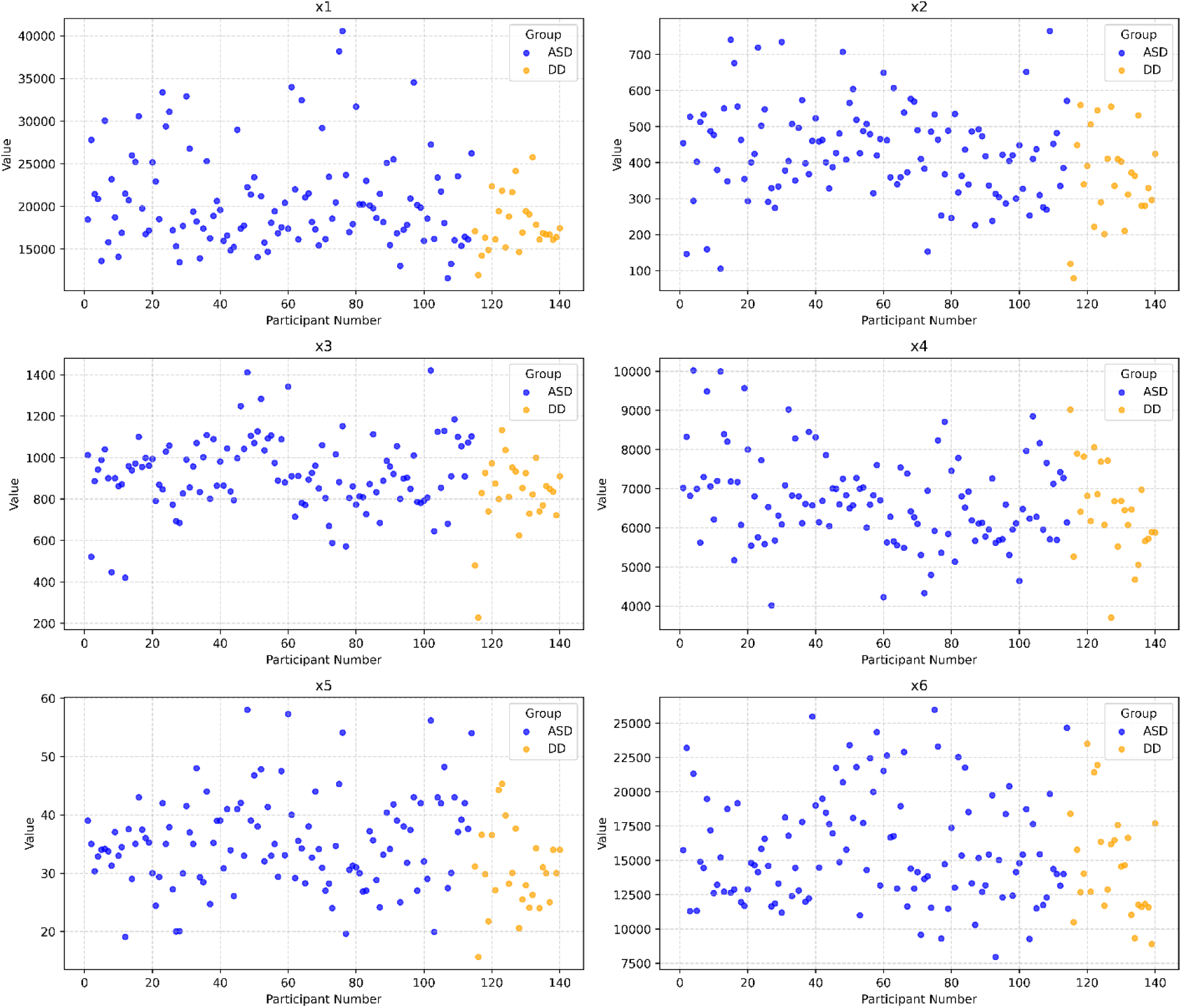
Scatterplots of data points of x1-x6.

**Fig. 6(b).**
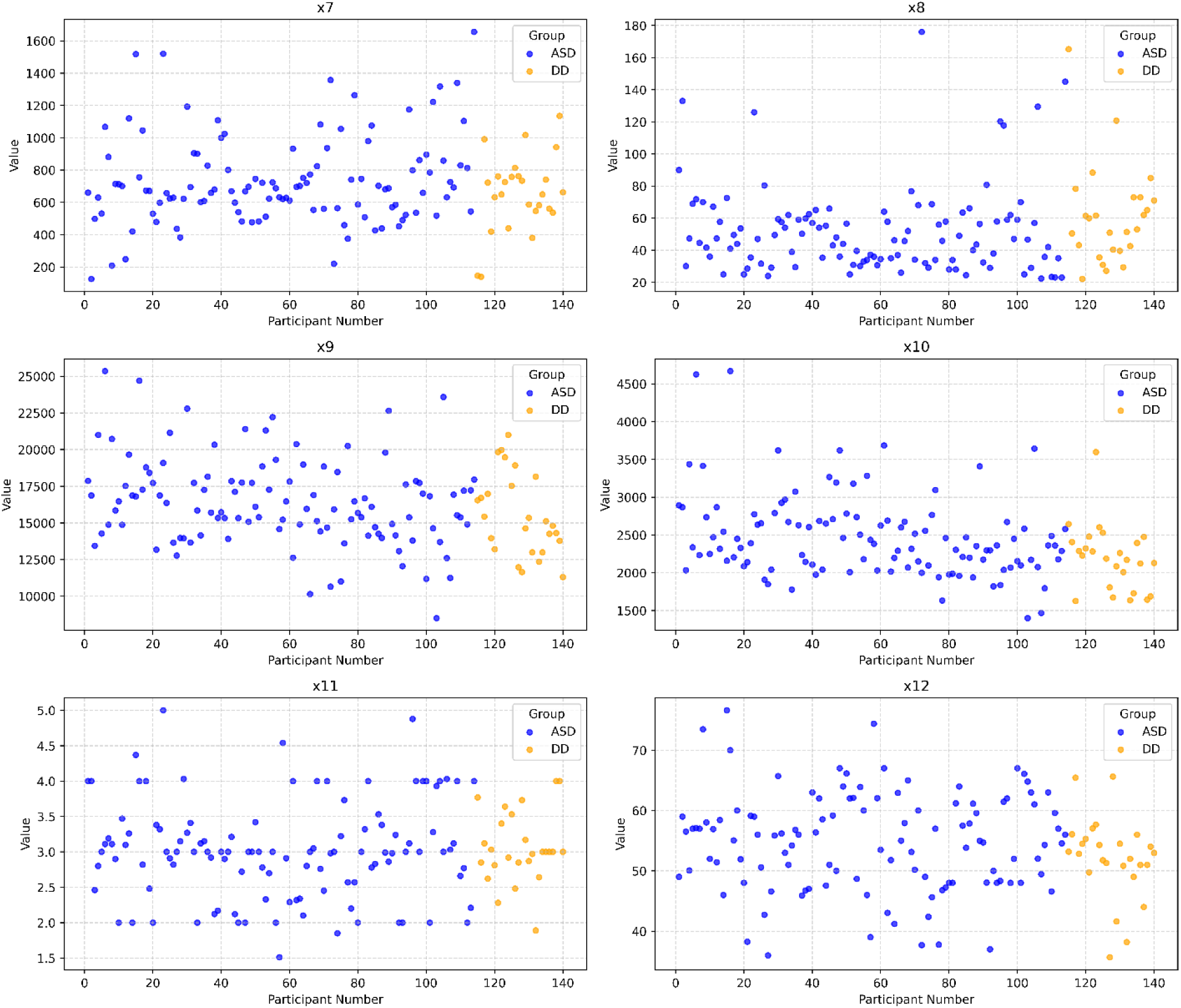
Scatterplots of data points of x7-x12.

**Fig. 6(c).**
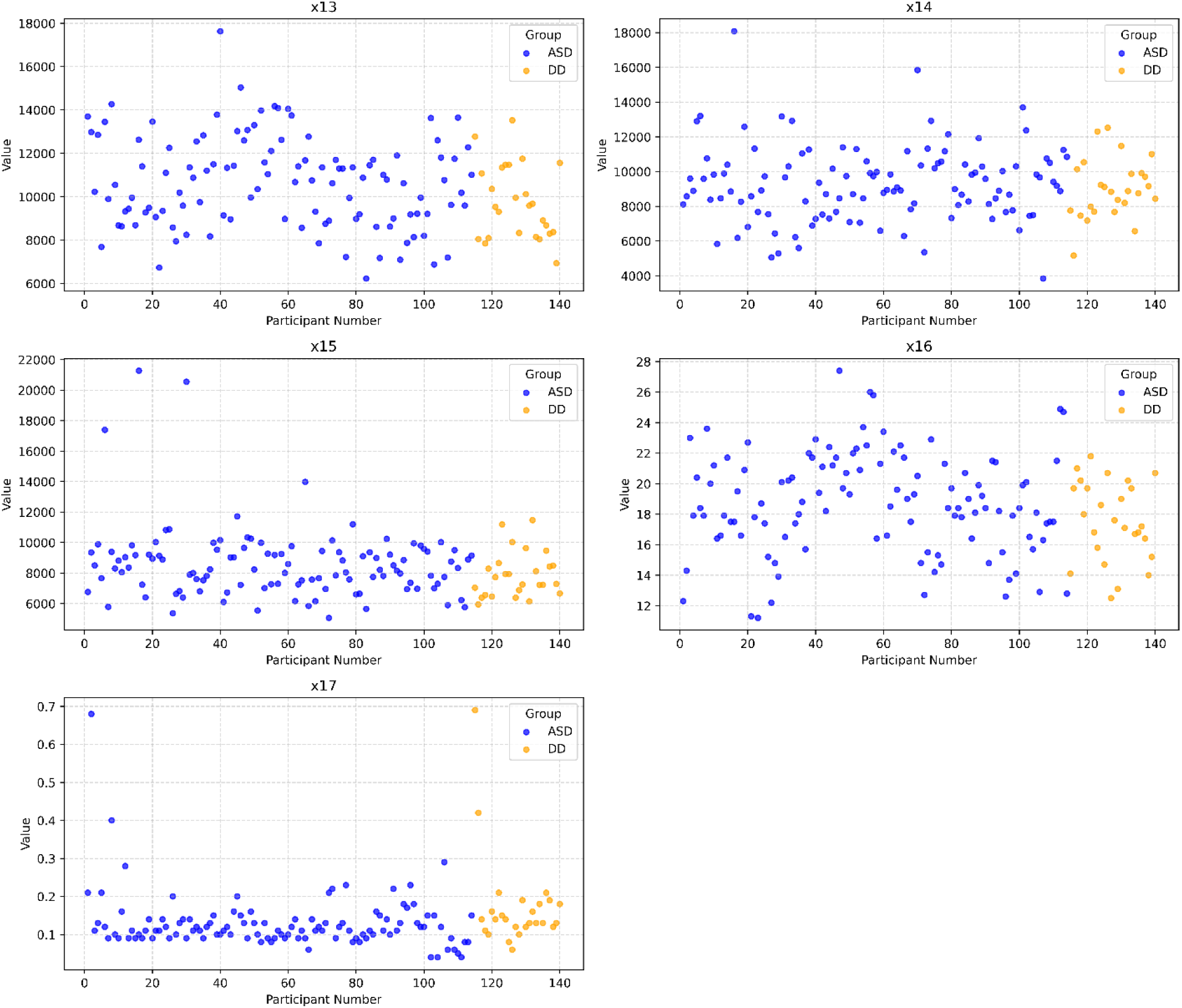
Scatterplots of data points of x13-x17.

**Table 6.**
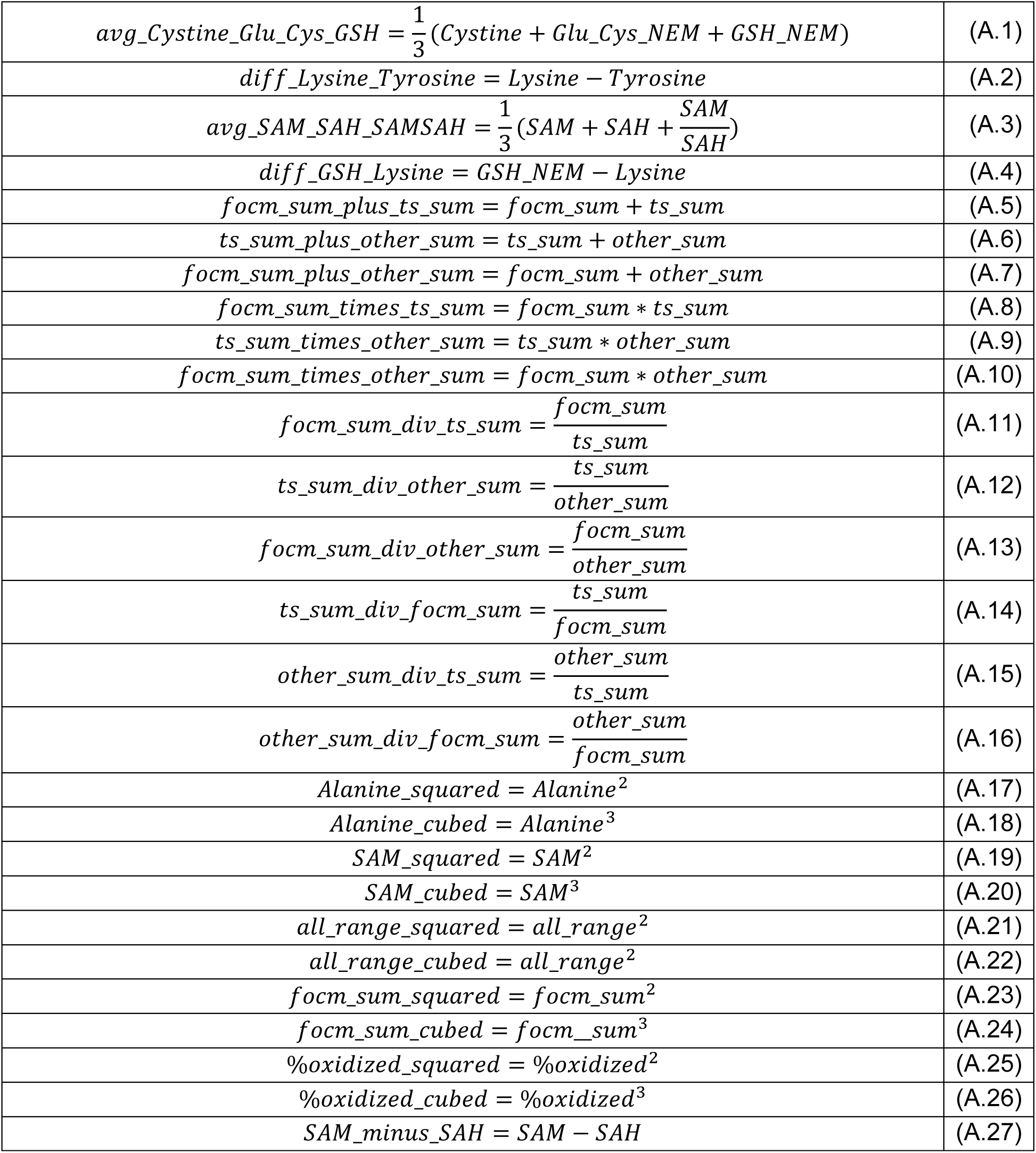
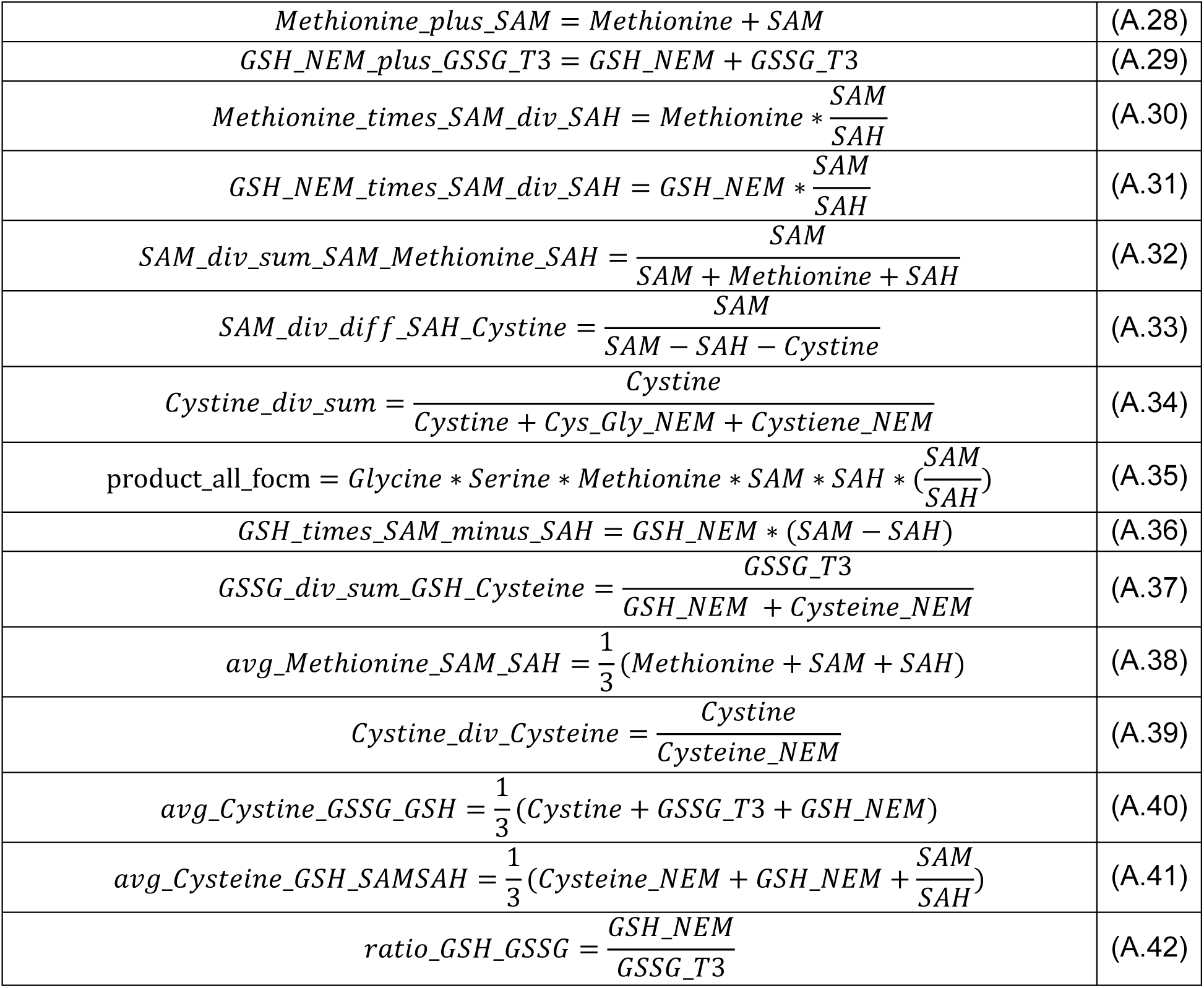
Equations of Engineered Features.

**Table 7.**
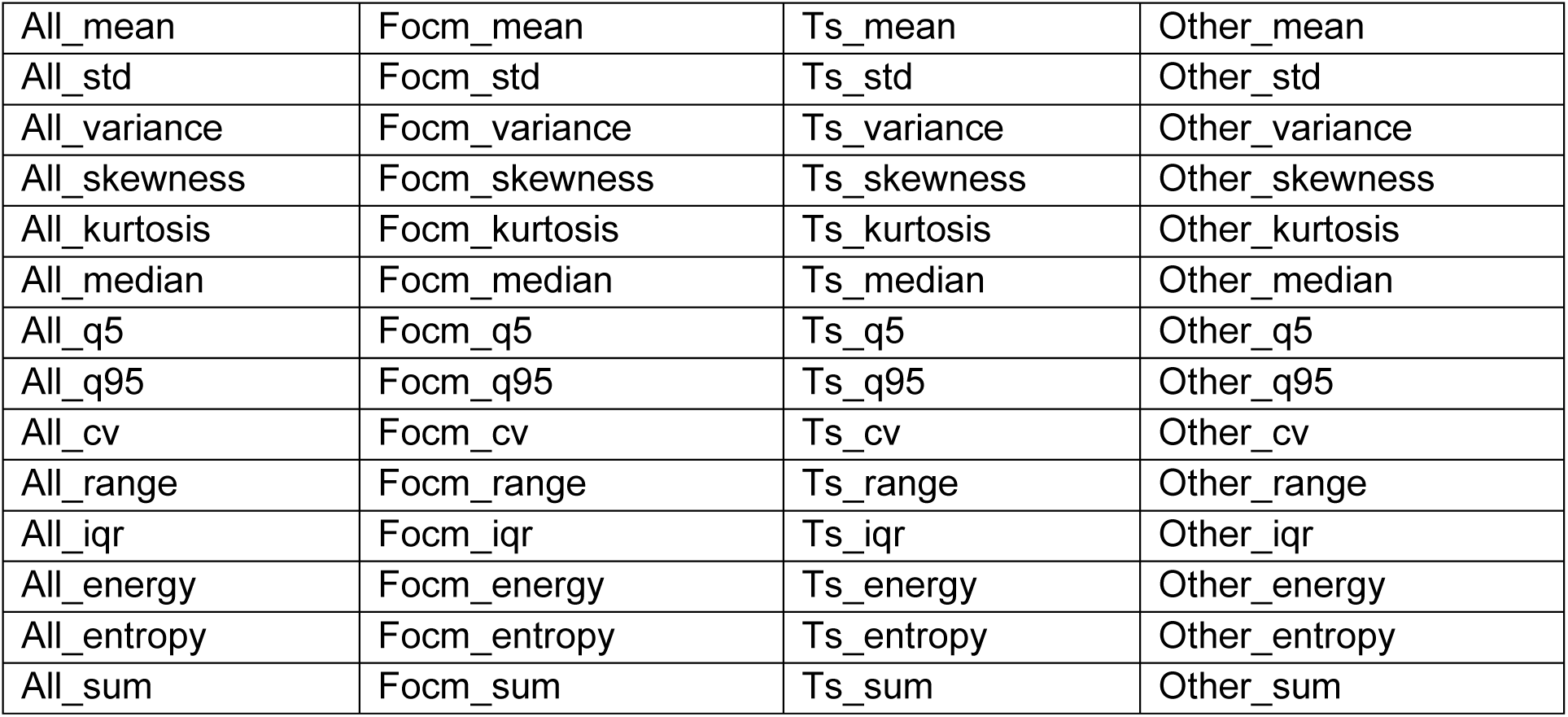
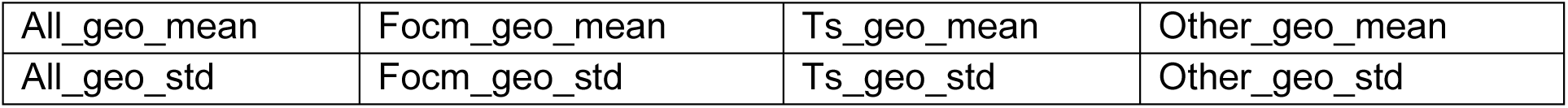
All Statistically Engineered Features.

|  |  |  |  |
| --- | --- | --- | --- |
| All_mean | Focm_mean | Ts_mean | Other_mean |
| All_std | Focm_std | Ts_std | Other_std |
| All_variance | Focm_variance | Ts_variance | Other_variance |
| All_skewness | Focm_skewness | Ts_skewness | Other_skewness |
| All_kurtosis | Focm_kurtosis | Ts_kurtosis | Other_kurtosis |
| All_median | Focm_median | Ts_median | Other_median |
| All_q5 | Focm_q5 | Ts_q5 | Other_q5 |
| All_q95 | Focm_q95 | Ts_q95 | Other_q95 |
| All_cv | Focm_cv | Ts_cv | Other_cv |
| All_range | Focm_range | Ts_range | Other_range |
| All_iqr | Focm_iqr | Ts_iqr | Other_iqr |
| All_energy | Focm_energy | Ts_energy | Other_energy |
| All_entropy | Focm_entropy | Ts_entropy | Other_entropy |
| All_sum | Focm_sum | Ts_sum | Other_sum |
| All_geo_mean | Focm_geo_mean | Ts_geo_mean | Other_geo_mean |
| All_geo_std | Focm_geo_std | Ts_geo_std | Other_geo_std |

**Table 8.** List of all Engineered Features from Equations in Table 6.

|  |  |
| --- | --- |
| focm_sum_plus_ts_sum | Alanine_squared |
| ts_sum_plus_other_sum | Alanine_cubed |
| focm_sum_plus_other_sum | SAM_squared |
| focm_sum_times_ts_sum | SAM_cubed |
| ts_sum_times_other_sum | all_range_squared |
| focm_sum_times_other_sum | all_range_cubed |
| focm_sum_div_ts_sum | focm_sum_squared |
| ts_sum_div_other_sum | focm_sum_cubed |
| focm_sum_div_other_sum | %oxidized_squared |
| ts_sum_div_focm_sum | %oxidized_cubed |
| other_sum_div_ts_sum | Methionine_times_SAM_div_SAH |
| other_sum_div_focm_sum | GSH_NEM_times_SAM_div_SAH |
| SAM_minus_SAH | SAM_div_sum_SAM_Methionine_SAH |
| Methionine_plus_SAM | SAM_div_diff_SAH_Cystine |
| GSH_NEM_plus_GSSG_T3 | avg_Cystine_Glu_Cys_GSH |
| Cystine_div_sum | GSSG_div_sum_GSH_Cysteine |
| product_all_focm | avg_Methionine_SAM_SAH |
| ratio_GSH_GSSG | GSH_times_SAM_minus_SAH |
| Cystine_div_Cysteine | avg_Cystine_GSSG_GSH |
| diff_Lysine_Tyrosine | avg_Cysteine_GSH_SAMSAH |
| avg_SAM_SAH_SAMSAH |  |
| diff_GSH_Lysine |  |

**Table 9.** Summary Statistics of the Metabolites Measured (Measurements are in ng/ml)

| Metabolite | ASD (n=114) Mean<br>± SD | ASD (n=114) Me-<br>dian [IQR] | DD (n=26) Mean<br>± SD | DD (n=26) Me-<br>dian [IQR] |
| --- | --- | --- | --- | --- |
| X1 | 20748.94 ±<br>5694.05 | 19425.00 [16875.00<br>- 23350.00] | 17863.94 ±<br>3179.68 | 16912.50<br>[16131.25 -<br>19356.25] |
| X2 | 428.99 ± 128.08 | 422.88 [339.19 -<br>500.62] | 354.15 ± 128.34 | 351.19 [282.44 -<br>420.62] |
| X3 | 924.43 ± 174.19 | 909.75 [816.50 -<br>1041.50] | 821.30 ± 179.59 | 842.00 [747.25 -<br>925.56] |
| X4 | 6700.23 ± 1154.04 | 6593.77 [5955.25 -<br>7270.12] | 6432.54 ±<br>1166.05 | 6432.09<br>[5758.50 -<br>6944.75] |
| X5 | 35.31 ± 7.89 | 34.84 [30.00 -<br>39.79] | 30.40 ± 7.04 | 30.00 [25.70 -<br>34.20] |
| X6 | 15822.80 ± 3993.49 | 14808.33 [12818.75 - 18512.50] | 14681.50 ± 3846.36 | 14293.75 [11718.75 - 16606.25] |
| X7 | 734.77 ± 276.68 | 684.21 [561.18 - 851.60] | 655.25 ± 235.36 | 656.50 [550.49 - 760.03] |
| X8 | 51.56 ± 26.77 | 46.44 [34.00 - 59.29] | 60.81 ± 30.72 | 56.44 [40.89 - 72.50] |
| X9 | 16419.80 ± 2993.35 | 16100.00 [14589.58 - 17843.75] | 15506.10 ± 2791.38 | 14956.50 [13343.75 - 17387.50] |
| X10 | 2472.37 ± 546.93 | 2361.50 [2100.50 - 2682.50] | 2193.10 ± 429.75 | 2207.50 [1857.50 - 2404.38] |
| X11 | 3.02 ± 0.70 | 3.00 [2.59 - 3.32] | 3.06 ± 0.50 | 3.00 [2.85 - 3.34] |
| X12 | 54.78 ± 8.18 | 55.45 [48.43 - 60.00] | 52.14 ± 6.75 | 52.91 [50.87 - 55.09] |
| X13 | 10660.00 ± 2104.53 | 10712.50 [9080.27 - 12058.25] | 9738.03 ± 1701.09 | 9558.75 [8306.00 - 11276.21] |
| X14 | 9285.66 ± 2186.32 | 9035.75 [7892.25 - 10405.62] | 8995.35 ± 1709.92 | 8858.75 [7806.88 - 9900.00] |
| X15 | 8596.02 ± 2393.62 | 8315.00 [7257.25 - 9384.38] | 7877.22 ± 1489.95 | 7511.50 [6721.00 - 8472.50] |
| X16 | 18.72 ± 3.32 | 18.45 [16.52 - 21.20] | 17.59 ± 2.62 | 17.40 [15.95 - 19.70] |
| X17 | 0.13 ± 0.07 | 0.11 [0.09 - 0.14] | 0.17 ± 0.12 | 0.14 [0.12 - 0.18] |

**Table 10.** Normality and Skewness Test Results.

| Feature | Group | Anderson-Darling (A) | Critical Value (5%) | Skewness | Skewness p-value | Assessment |
| --- | --- | --- | --- | --- | --- | --- |
| X1 | ASD (n=114) | 3.47 | 0.76 | 1.2 | <0.001 | Non-normal, Right-skewed |
|  | DD (n=26) | 0.79 | 0.7 | 0.82 | 0.073 | Non-normal, Symmetric |
| X2 | ASD (n=114) | 0.35 | 0.76 | 0.21 | 0.336 | Normal, Symmetric |
|  | DD (n=26) | 0.21 | 0.7 | -0.2 | 0.644 | Normal, Symmetric |
| X3 | ASD (n=114) | 0.58 | 0.76 | -0.02 | 0.942 | Normal, Symmetric |
|  | DD (n=26) | 0.9 | 0.7 | -1.52 | 0.003 | Non-normal, Left-skewed |
| X4 | ASD (n=114) | 0.9 | 0.76 | 0.53 | 0.021 | Non-normal, Right-skewed |
|  | DD (n=26) | 0.23 | 0.7 | 0 | 0.997 | Normal, Symmetric |
| X5 | ASD (n=114) | 0.62 | 0.76 | 0.51 | 0.027 | Normal, Right-skewed |
|  | DD (n=26) | 0.21 | 0.7 | 0.26 | 0.55 | Normal, Symmetric |
| X6 | ASD (n=114) | 1.98 | 0.76 | 0.61 | 0.009 | Non-normal, Right-skewed |
|  | DD (n=26) | 0.49 | 0.7 | 0.67 | 0.136 | Normal, Symmetric |
| X7 | ASD (n=114) | 2.64 | 0.76 | 0.93 | <0.001 | Non-normal, Right-skewed |
|  | DD (n=26) | 0.4 | 0.7 | -0.3 | 0.493 | Normal, Symmetric |
| X8 | ASD (n=114) | 5.47 | 0.76 | 2.12 | <0.001 | Non-normal, Right-skewed |
|  | DD (n=26) | 0.96 | 0.7 | 1.79 | <0.001 | Non-normal, Right-skewed |
| X9 | ASD (n=114) | 0.79 | 0.76 | 0.43 | 0.061 | Non-normal, Symmetric |
|  | DD (n=26) | 0.4 | 0.7 | 0.41 | 0.347 | Normal, Symmetric |
| X10 | ASD (n=114) | 2.66 | 0.76 | 1.41 | <0.001 | Non-normal, Right-skewed |
|  | DD (n=26) | 0.67 | 0.7 | 1.13 | 0.018 | Normal, Right-skewed |
| X11 | ASD (n=114) | 1.96 | 0.76 | 0.35 | 0.116 | Non-normal, Symmetric |
|  | DD (n=26) | 0.64 | 0.7 | 0.06 | 0.889 | Normal, Symmetric |
| X12 | ASD (n=114) | 0.32 | 0.76 | 0 | 0.993 | Normal, Symmetric |
|  | DD (n=26) | 1.02 | 0.7 | -0.51 | 0.245 | Non-normal, Symmetric |
| X13 | ASD (n=114) | 0.46 | 0.76 | 0.3 | 0.182 | Normal, Symmetric |
|  | DD (n=26) | 0.56 | 0.7 | 0.5 | 0.258 | Normal, Symmetric |
| X14 | ASD (n=114) | 0.58 | 0.76 | 0.67 | 0.005 | Normal, Right-skewed |
|  | DD (n=26) | 0.22 | 0.7 | 0.19 | 0.664 | Normal, Symmetric |
| X15 | ASD (n=114) | 4.96 | 0.76 | 2.83 | <0.001 | Non-normal, Right-skewed |
|  | DD (n=26) | 0.7 | 0.7 | 0.98 | 0.036 | Normal, Right-skewed |
| X16 | ASD (n=114) | 0.25 | 0.76 | -0.06 | 0.801 | Normal, Symmetric |
|  | DD (n=26) | 0.34 | 0.7 | -0.28 | 0.518 | Normal, Symmetric |
| X17 | ASD (n=114) | 9.19 | 0.76 | 4.41 | <0.001 | Non-normal, Right-skewed |
|  | DD (n=26) | 3.65 | 0.7 | 3.36 | <0.001 | Non-normal, Right-skewed |

**Table 11.** Confidence Intervals (95%) of the Metrics Reported (Monte Carlo Simulations)

| No of Features | Balanced Accuracy | AUC | Accuracy | Sensitivity | Specificity | Precision | F1 | MCC |
| --- | --- | --- | --- | --- | --- | --- | --- | --- |
| 4 | (0.5565, 0.9783) | (0.6435, 1) | (0.75, 0.9643) | (0.7826, 1) | (0.2, 1) | (0.84, 1) | (0.8444, 0.9787) | (0.1400, 0.8928) |
| 5 | (0.5783, 0.9783) | (0.6522, 1) | (0.7857, 0.9643) | (0.8261, 1) | (0.2, 1) | (0.8462, 1) | (0.8571, 0.9787) | (0.5674, 0.9627) |

